# Emerging SARS-CoV-2 variants of concern evade humoral immune responses from infection and vaccination

**DOI:** 10.1101/2021.05.26.21257441

**Authors:** Tom G. Caniels, Ilja Bontjer, Karlijn van der Straten, Meliawati Poniman, Judith A. Burger, Brent Appelman, Ayesha H.A. Lavell, Melissa Oomen, Gert-Jan Godeke, Coralie Valle, Ramona Mögling, Hugo D.G. van Willigen, Elke Wynberg, Michiel Schinkel, Lonneke A. van Vught, Denise Guerra, Jonne L. Snitselaar, Devidas N. Chaturbhuj, Isabel Cuella Martin, Amsterdam UMC COVID-19 S3/HCW study group, John P. Moore, Menno D. de Jong, Chantal Reusken, Jonne J. Sikkens, Marije K. Bomers, Godelieve J. de Bree, Marit J. van Gils, Dirk Eggink, Rogier W. Sanders

**Author notes:** These authors contributed equally to this work (Tom G. Caniels, Ilja Bontjer, Karlijn van der Straten).

## Abstract

Emerging SARS-CoV-2 variants pose a threat to human immunity induced by natural infection and vaccination. We assessed the recognition of three variants of concern (B.1.1.7, B.1.351 and P.1) in cohorts of COVID-19 patients ranging in disease severity (n = 69) and recipients of the Pfizer/BioNTech vaccine (n = 50). Spike binding and neutralization against all three VOC was substantially reduced in the majority of samples, with the largest 4-7-fold reduction in neutralization being observed against B.1.351. While hospitalized COVID-19 patients and vaccinees maintained sufficient neutralizing titers against all three VOC, 39% of non-hospitalized patients did not neutralize B.1.351. Moreover, monoclonal neutralizing antibodies (NAbs) show sharp reductions in their binding kinetics and neutralizing potential to B.1.351 and P.1, but not to B.1.1.7. These data have implications for the degree to which pre-existing immunity can protect against subsequent infection with VOC and informs policy makers of susceptibility to globally circulating SARS-CoV-2 VOC.

## Introduction

With over 140 million confirmed infections and over three million deaths as of April 2021, the coronavirus disease 2019 (COVID-19) pandemic shows few signs of abating (*1*). While severe acute respiratory syndrome coronavirus 2 (SARS-CoV-2), the causative agent of COVID-19, remained relatively stable genetically and antigenically during the first stage of the pandemic, higher levels of genetic variation have been observed during the second wave of the pandemic with considerable genetic changes compared to the original Wuhan Hu-1 strain of SARS-CoV-2. These genetic changes include substitutions within the functional domains of the SARS-CoV-2 spike (S) protein resulting in altered phenotypes of the virus. The WHO has currently defined three VOC based on possible increase in transmissibility or change in COVID-19 epidemiology, increase in virulence or change in clinical disease presentation, or decrease in effectiveness of available diagnostics, vaccines and therapeutics. These VOC include B.1.1.7 (20I/N501Y.V1, first detected in the United Kingdom), B.1.351 (20H/N501Y.V2, first detected in South Africa) and B.1.1.28.P1 (P.1, 20J/N501Y.V3, first detected in Brazil), which have since spread globally and as of April 2021, cases with these variants have been reported in 132, 82 and 52 countries, respectively (*2*–*5*). The emergence of these variants has raised concerns as to whether immunity, be it natural immunity from prior infection or immunity from vaccination, can protect against these different variants. Moreover, it is unclear whether therapeutic monoclonal NAbs isolated from convalescent donors retain therapeutic efficacy against these new variants.

The S protein of coronaviruses is the main target of NAbs and consists of a membrane-proximal S2 domain containing the fusion peptide and a membrane-distal S1 domain containing the receptor binding domain (RBD) and the N-terminal domain (NTD) (Fig. 1A). The S protein mediates viral entry through interaction with the angiotensin-converting enzyme 2 (ACE2) receptor on host cells (*6, 7*). Therefore, substitutions present in the RBD of these emerging VOC are particularly worrisome as they might improve binding to the human receptor and thus increase viral fitness and transmissibility. Indeed, B.1.1.7, B.1.351 and P.1 all share the N501Y RBD mutation, which contributes to an enhanced interaction with the human ACE2 receptor resulting in increased infectivity and transmissibility (Fig. 1A) (*8, 9*). In addition, both B.1.351 and P.1 contain the E484K substitution and a substitution at position 417 (K417N in B.1.351 and K417T in P.1) both of which have been implicated in escape from NAbs by several studies (Fig. 1A) (*10, 11*). More recently, E484K has also been observed in B.1.1.7 variants that have adopted this mutation independently, leading to a more substantial loss of neutralizing titers in vaccinated individuals than for B.1.1.7 alone (*12*). Additional substitutions mostly arise in the NTD region, located apically on the S1 domain. Although the exact functional implication of substitutions within the NTD is not clear, it has been shown that this domain is an important target for NAbs (*13, 14*). VOC B.1.351 and P.1 both carry five mutations in this region of which they share one (L18F). Early reports on the B.1.351 lineage reported variants with a substitution at position 242 and 246 (L242H and R246I), while other sublineages had a three amino acid deletion (Δ242-244) (*4*). B.1.1.7 does not have substitutions in its NTD but has deletions in this region (Δ69-70, Δ144) (Fig. 1A). Other mutations are identified in the S2 domain of the S protein and their impact on antibody recognition and infectivity is not yet well understood, partly because the vast majority of NAbs target the RBD or NTD. Finally, all these variants have the D614G mutation that defines the B.1 lineage and became dominant throughout 2020, now being present in the large majority of sequenced SARS-CoV-2 variants (> 99%) (Fig. 1A) (*15*).

**Figure 1.**
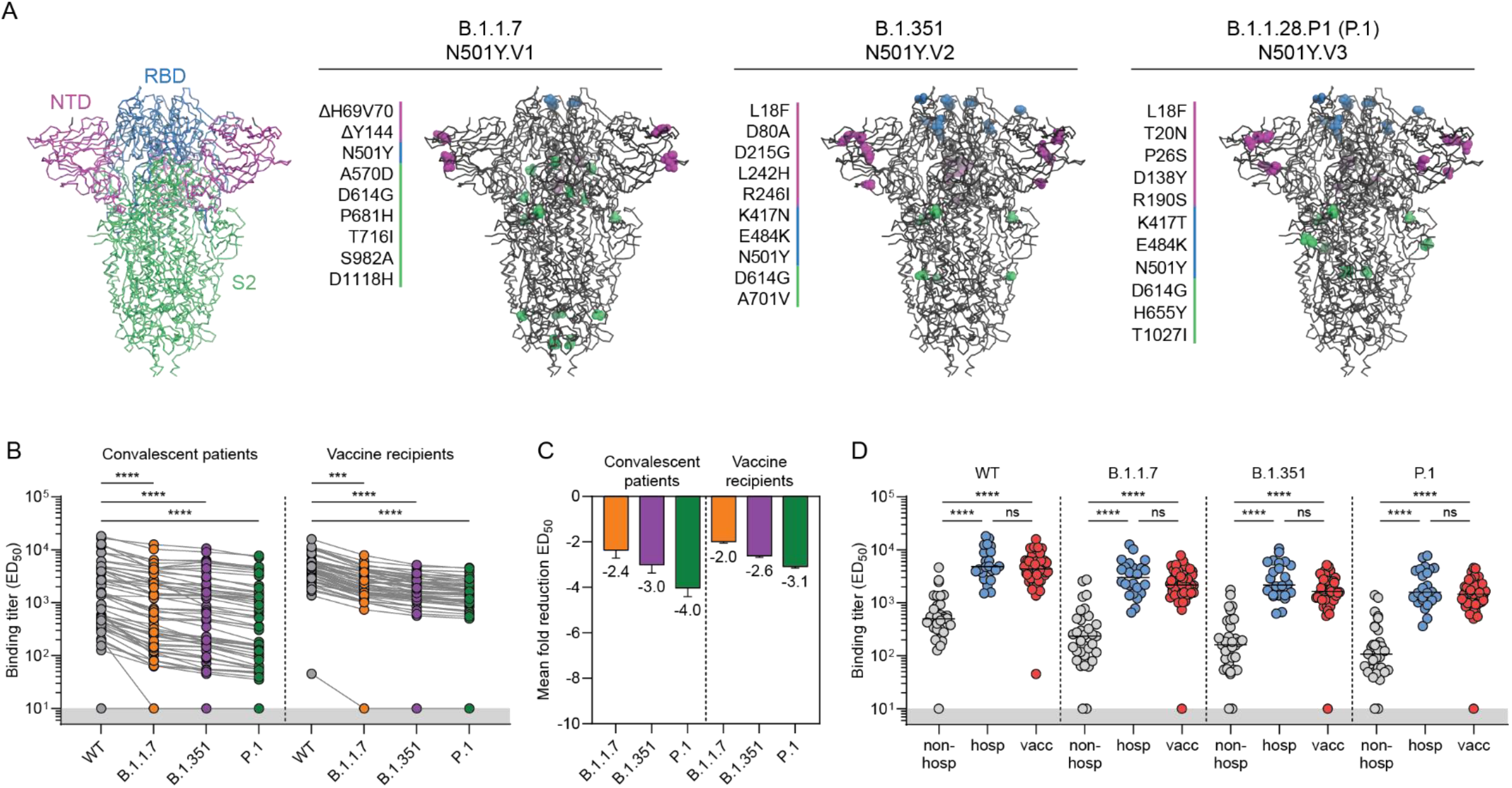
Binding of convalescent and vaccinee sera to VOC B.1.1.7, B.1.351 and P.1 spike proteins. (**A**) Structural representation of spike (S) protein with its three domains (N-terminal domain, NTD, in magenta; receptor-binding domain (RBD), in blue; S2 domain in green). Mutations in each of the VOC S proteins are listed as well as their location in the trimer. Colors correspond to the S protein domains in which the mutation occurs in. (**B**) Half-maximal binding (ED_50_) titers of polyclonal convalescent sera (left, n = 57) and vaccinee sera (right, n = 50) to S protein of B.1.1.7, B.1.351 and P.1 VOC. Connected dots indicate results from the same individual. The lower cutoff for binding was set at an ED_50_ of 10 (grey shading). (**C**) Mean ± SEM fold reductions in ED_50_ titers for convalescent patients and vaccine recipients against S proteins of B.1.1.7, B.1.351 and P.1 VOC in comparison to ED_50_ titers to the WT S protein. (**D**) ED_50_ titers of non-hospitalized COVID-19 patients, hospitalized COVID-19 patients and vaccine recipients against WT S protein and each of the VOC S protein. ****, *p* < 0.0001; ***, *p* < 0.001; ns, not significant. All data points shown here represent the mean of a technical triplicate.

All three VOC have been linked to increased infectivity and transmissibility, and the first reports about specific substitutions in B.1.351 and P.1 associated with escape from immunity by infection or mRNA vaccines have become available (*10, 16*–*18*). These first reports suggest that pre-existing immunity was generally sufficient to neutralize B.1.1.7 to similar levels as WT in mRNA vaccine recipients and in convalescent individuals up to nine months after infection (*17, 19*). In case of B.1.351, the impact is more substantial with neutralizing titers from convalescent patients being reduced by ∼6 to 13-fold while vaccine recipients are reported to have a ∼10 to 14-fold reduction (*10, 17, 18*). Moreover, it is reported that ∼40% of convalescent patients do not have any neutralizing activity against B.1.351 nine months after primary infection (*17*). A recent study showed that although B.1.351 and P.1 have similar mutations in their RBD, sera from convalescent patients as well as mRNA vaccine recipients showed a neutralization reduction of ∼3-fold against P.1 while this was 7-13-fold for B.1.351 (*20, 21*).

Although these studies have provided valuable initial insights in altered antigenic properties of these VOC, few studies have compared all three VOC side-by-side. Furthermore, comparing immune responses from vaccine recipients and individuals who have suffered from mild or severe COVID-19 as well as reactivity of NAbs to all three VOC within the same study will provide useful insights that can be used for additional surveillance of SARS-CoV-2 variants, the use of monoclonal antibodies for treatment and modifications of vaccines in order to increase immune coverage of immunity to include yet unknown emerging SARS-CoV-2 variants.

## Results

### Recognition of SARS-CoV-2 VOC by convalescent and vaccine sera is reduced

Here, we assessed the impact of VOC B.1.1.7, B.1.351 and P.1 on humoral immunity elicited either by mRNA vaccination or by natural infection with SARS-CoV-2. To this end, we studied two cohorts of individuals: the COSCA cohort that included convalescent COVID-19 patients (n = 69) and the S3 cohort that included health care workers (HCW) who were vaccinated twice with the Pfizer/BioNTech COVID-19 vaccine (n = 50) (Table 1). The COSCA cohort included hospitalized and non-hospitalized patients in the Netherlands who were enrolled and sampled four to six weeks after symptom onset (i.e. close to the expected peak of humoral immunity) between March 2020 and January 2021 (Table 1). Based on the rarity of B.1.351 and P.1 in the Netherlands, no individuals are expected to have been infected with either VOC. However, infections with B.1.1.7 became more prevalent towards the end of 2020, with the estimated prevalence of B.1.1.7 reaching 24% in the Netherlands in the week of the last inclusion (January 2021) (*22*). As genomic data was not collected as part of this study, we cannot exclude that a few patients were infected with B.1.1.7. The S3 cohort consists of health care workers without prior SARS-CoV-2 infection who received the approved Pfizer/BioNTech COVID-19 mRNA vaccine twice with a three week interval and sampled four weeks after the second vaccination (Table 1).

**Table 1.**
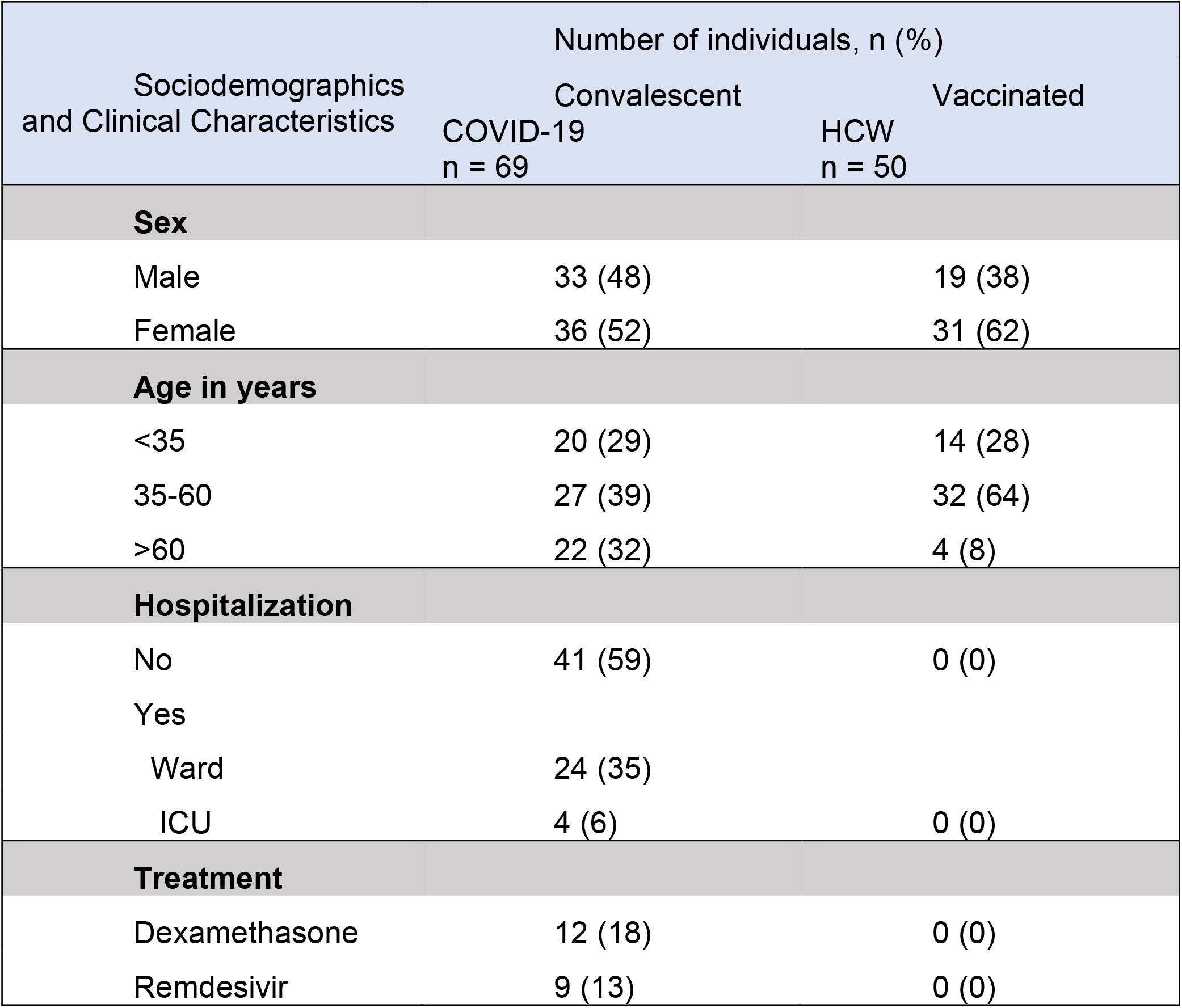
Sociodemographic and clinical characteristics of study populations.

We first assessed S protein binding titers of convalescent and vaccinee sera in a custom multiplex protein microarray that has been extensively validated for clinical use (table S1) (*23*). We generated S proteins, using previously described stabilization approaches, from all three VOC that had been identified at the time of assessment as well as a control wild-type (WT) S protein from the Wuhan Hu-1 virus (GenBank: MN908947.3) isolated in December 2019 (*6, 24, 25*). Overall, the antibody responses against each S protein were heterogeneous and differed up to ∼1200-fold between the strongest and weakest responders. The recognition of the three VOC by convalescent patients was significantly reduced compared to WT by an average of 2.4-fold, 3-fold and 4-fold for B.1.1.7, B.1.351 and P.1, respectively (*p* < 0.0001 for all, Fig. 1, B and C). Binding titers elicited by the mRNA vaccine were more homogeneous than those elicited by natural infection and differed ∼10-fold between responders, with all participants having half-maximal binding titers (ED_50_s) exceeding 10^3^, except for one poor responder (Fig. 1B). Since the variability of binding titers in convalescent sera is considerable, we examined whether this variability was related to severity of disease (i.e. hospital admission, Fig. 1D). We observed a highly significant ∼8-fold difference in ED_50_s between non-hospitalized patients and hospitalized patients, which is in line with previous reports of WT SARS-CoV-2 binding titers correlating with severity of disease (*p* < 0.0001) (*26, 27*). This difference in binding titers between non-hospitalized patients and hospitalized patients was consistent for all three SARS-CoV-2 VOC studied (Fig. 1D). When comparing immune responses of vaccine recipients with convalescent sera, we observed similar S protein binding titers between vaccinee and hospitalized patients, which are an average ∼4 to 11-fold higher compared to non-hospitalized patients for all VOC (Fig. 1D). Taken together, these data indicate that vaccine recipients as well as COVID-19 patients exhibit reduced binding to S proteins of the currently circulating VOC, with hospitalized patients and vaccine recipients exhibiting higher binding titers overall compared to non-hospitalized patients.

### SARS-CoV-2 VOC are substantially less sensitive to serum NAbs

We next tested the neutralizing activity of convalescent and vaccinee sera (Fig. 2, table S1). To this end, we generated lentiviral-based pseudoviruses of the currently widespread SARS-CoV-2 D614G (WT) variant as well as B.1.1.7, B.1.351 and P.1. We detected substantial neutralizing activity against the WT virus (half maximal neutralization titer, ID_50_ > 100) in 96% of convalescent patients irrespective of hospitalization, and in all vaccine recipients, except the one poor responder (Fig. 2A). Indeed, the overall binding titers correlated well with the neutralization titers for all VOC (fig. S1C). Non-hospitalized patients had the most heterogeneous responses, with WT neutralizing titers differing by up to ∼150-fold, while titers against WT were much more homogeneous in hospitalized patients (up to ∼20-fold difference) and vaccine recipients (up to ∼12-fold difference) (Fig. 2A). Consistent with the binding results, we observed a marked and significant reduction in serum ability to neutralize VOC pseudoviruses (Fig. 2A). For all three groups, the difference was most apparent against the B.1.351 VOC, showing a reduction of ∼4-, 7- and 5- fold in neutralizing titers for non-hospitalized patients, hospitalized patients and vaccine recipients, respectively (*p* < 0.0001, Fig. 2, A and B). The data were corroborated in an authentic SARS-CoV-2 neutralization assay (fig. S2, A-D). We did not observe an effect of medication, including the antiviral drug remdesivir (rdv) and the anti-inflammatory drug dexamethasone (dexa) on neutralization activity in sera from convalescent but previously hospitalized patients (fig. S2E).

**Figure 2.**
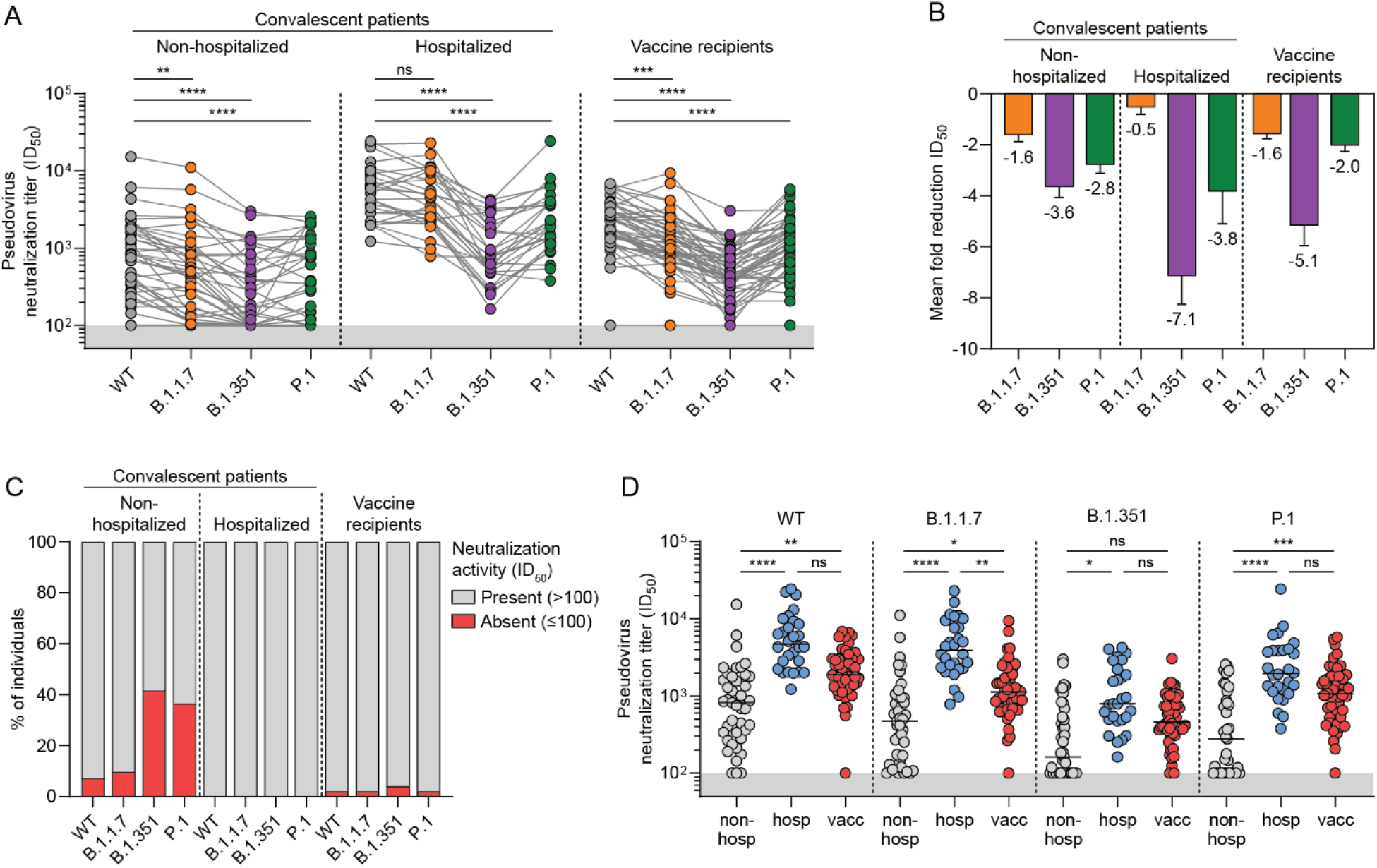
Neutralization of VOC B.1.1.7, B.1.351 and P.1 by convalescent and vaccinee sera. (**A**) Half-maximal neutralization (ID_50_) titers of polyclonal sera from non-hospitalized convalescent patients (n = 41), hospitalized convalescent patients (n = 28) and vaccine recipients (n = 50) against pseudoviruses of WT and B.1.1.7, B.1.351 and P.1 VOC. Connected dots indicate results from the same individual. The lower cutoff for neutralization was set at an ID_50_ of 100 (grey shading). (**B**) Mean ± SEM fold reductions in ID_50_ titers for non-hospitalized convalescent patients, hospitalized convalescent patients and vaccine recipients against B.1.1.7, B.1.351 and P.1 VOC pseudoviruses in comparison to ID_50_ titers against the WT pseudovirus. (**C**) Percentage of individuals in each of the three groups in (**B**) that have no detectable serum neutralizing activity (ID_50_ < 100) against the indicated pseudoviruses. (**D**) ID_50_ titers of non-hospitalized patients, hospitalized patients and vaccine recipients against WT pseudovirus and each of the VOC pseudoviruses. ****, *p* < 0.0001; ***, *p* < 0.001; **, *p* < 0.01; *, *p* < 0.05; ns, not significant. All data points shown here represent the mean of a technical triplicate and are representative of at least two independent experiments.

Some sera, most notably those from non-hospitalized patients, showed a complete loss of neutralization against some VOC, whereas they did neutralize WT pseudovirus (Fig. 2A). When the sera that reached the limit of detection against one of the VOC (i.e., ID_50_ < 100) were excluded, the fold difference observed between WT and VOC neutralization remained very similar across the three groups (fig. S2F). 39% (16/41) non-hospitalized patients in this study lost all neutralizing activity against B.1.351 and 34% (14/41) were unable to neutralize P.1 while they did neutralize WT pseudovirus and B.1.1.7 (Fig. 2C). In contrast, all hospitalized patients (28/28) retained at least some neutralizing activity against B.1.1.7, B.1.351 and P.1 and only 1/50 vaccine recipients completely lost neutralizing activity against B.1.351, suggesting that high titers of NAbs against WT are predictive for cross-neutralization of VOC (Fig. 2C). Neutralizing titers of vaccine recipients against B.1.1.7 were an average ∼3.6-fold lower than those of hospitalized patients (*p* = 0.0015), while this was less substantial and not significant for B.1.351 (∼2.2-fold) and P.1 (∼2.4-fold, Fig. 2D). Although B.1.351 showed the largest reduction in serum neutralization titers, the largest reduction in binding for both convalescent patients and vaccinee was seen against P.1 (Fig. 1C and Fig. 2B). Overall, we conclude that high neutralization levels against WT virus are predictive for the ability to neutralize VOC, while low neutralization levels against WT often translate to the inability to neutralize VOC B.1.351 and P.1.

Although hospitalized patients did not show a significant reduction in B.1.1.7 neutralization, both vaccine recipients and non-hospitalized patients showed a small but statistically significant 1.6-fold reduction for B.1.1.7 compared to WT (*p* = 0.0035, Fig. 2, A and B). The sole RBD mutation in B.1.1.7, N501Y, might contribute to this effect although it has only been sporadically implicated in NAb escape (*28*). Alternatively, the two NTD deletions in B.1.1.7 (Δ69-70, Δ144, Fig. 1A) that are not present in WT, B.1.351 or P.1 might play a role. Moreover, while P.1 carries amino acid substitutions in the RBD at the exact same positions as B.1.351, the reduction in P.1 neutralization compared to WT was significantly less than for B.1.351 (*p* < 0.001, Fig. 2, A and B), which might be explained by differences in the NTD.

### Neutralizing monoclonal NAbs lose potency against VOC

To obtain more insight into the antibody specificities that were affected by the mutations in the VOC we assessed a panel of NAbs with known specificities isolated from convalescent patients, some of which have since been characterized structurally, and some of which have been evaluated in preclinical protection models (*25, 29, 30*). Bio-layer interferometry (BLI) experiments showed that for the majority of RBD-targeting NAbs that span the four known epitope clusters on the RBD (*31*) the binding to B.1.351 and P.1 S protein was reduced substantially, while binding to the B.1.1.7 S protein was mostly similar to WT binding. This is consistent with observations that the only RBD mutation in B.1.1.7, i.e. N501Y, has not been associated with escape from antibodies and was probably selected for increased ACE2 binding (*8, 9*), while E484K and K417N/T have a larger impact in neutralization by NAbs. RBD-targeting NAb COVA1-18, which neutralized WT with a half-maximal inhibitory concentration (IC_50_) in the ng/mL range and protected from WT infections in three preclinical animal models (*25, 30*), displayed dramatically reduced binding to B.1.351 and P.1 S proteins (Fig. 3A). Similarly, COVA2-15, which is highly potent against WT, showed decreased binding kinetics to S proteins from B.1.351 and P.1 compared to WT and B.1.1.7 S proteins (Fig. 3A). We also tested binding of RBD-targeting, SARS-CoV cross-NAbs COVA1-16 and COVA2-02, which showed highly similar binding to all VOC S proteins tested, indicating that these NAbs in particular target a conserved epitope that has not been influenced by the mutations present in these three VOC. This is in line with previous findings of COVA1-16 able to recognize RBDs of pangolin and bat origin, indicating that COVA1-16 recognizes an epitope that is highly conserved among sarbecoviruses (*29*). For the NTD-targeting NAbs COVA1-22 and COVA 2-17, we found a substantial reduction of binding to B.1.351 and P.1. COVA1-22 also showed reduced binding to B.1.1.7 S protein, while COVA2-17 retained binding to B.1.1.7. These results show that the binding of RBD NAbs as well as NTD NAbs are impacted by mutations in VOC, while there are some RBD NAbs that retain similar binding kinetics to all VOC.

**Figure 3.**
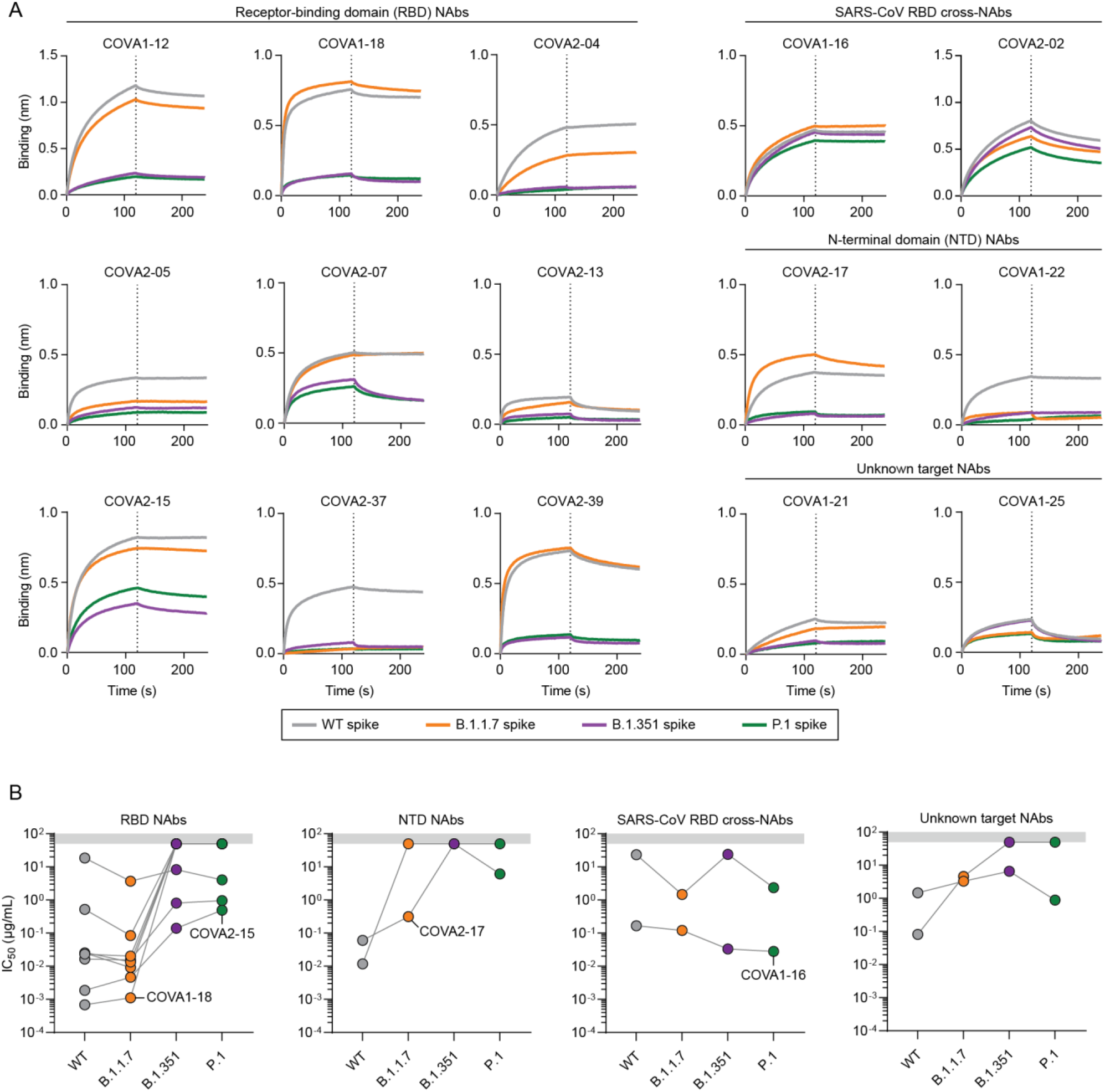
Binding kinetics and neutralization of NAbs isolated from convalescent COVID-19 patients. (**A**) Bio-layer interferometry (BLI) sensorgrams of 15 NAbs binding profile to WT spike as well as to each of the VOC S proteins (WT, grey; B.1.1.7, orange; B.1.351, purple; P.1, green). The dotted line indicates the end of NAb association and the start of dissociation. (**B**) Half-maximal inhibitory concentrations (IC_50_) values of 15 NAbs to each of the pseudoviruses used in Figure 2., separated by their target epitope on S protein. COVA1-18, COVA2-15, COVA2-17 and COVA1-16 are highlighted because of their neutralization characteristics. The grey shading indicates the maximum NAb concentration tested (50 µg/mL). Connected dots indicate IC_50_ values from the same NAb. Each dot represents the mean ± SD of a technical triplicate. The curves shown are representative of at least two independent experiments.

Next, we attempted to pinpoint which mutations have the most impact on binding of RBD NAbs. Therefore, we expressed soluble RBDs of VOC B.1.1.7, B.1.351 and P.1 as well as a RBD with only the E484K mutation and performed an enzyme-linked immunosorbent assay (ELISA) with our panel of RBD NAbs (fig. S3, A and B). Similar to our BLI data (Fig. 3A), most RBD antibodies show a large and similar reduction in binding to B.1.351 and P.1 RBDs. However, COVA2-15 shows a ∼4-fold lower half-maximal binding titer (EC_50_) to P.1 compared to B.1.351, indicating that the K417N mutation impacts binding of this particular highly potent antibody more than K417T (*25*) (fig. S3, A and B). A similar observation can be made for COVA2-07, which does retain some binding against P.1 RBD but not against B.1.351 RBD (fig. S3, A and B). Interestingly, while E484K alone knocks out binding in COVA1-12, COVA1-18, COVA2-29 and COVA2-39, this is not the case for the other RBD NAbs that are not impacted by E484K unless it is accompanied by N501Y and a substitution at position 417 (fig. S3, A and B). For SARS-CoV cross-NAbs COVA1-16, COVA2-02 and CR3022, none of the substitutions found in the VOC studied here had any effect on binding. Thus, we show that while E484K alone has a substantial effect on the binding of some NAbs, it is the combination of E484K with other mutations such as K417N/T that have the greatest impact on the binding of RBD NAbs.

Finally, we sought to confirm these findings in pseudovirus neutralization assays (Fig. 3B, fig. S4). In line with previous results (*25*), COVA1-18 and COVA2-15 exhibit picomolar IC_50_ titers against WT and B.1.1.7 while neutralization against B.1.351 was completely knocked out for COVA1-18 and reduced by ∼100-fold for COVA2-15 (Fig. 3B, fig. S4). Of note, only 5/11 tested RBD NAbs retained any neutralization against B.1.351 and P.1, consistent with their abilities to bind the S proteins of these VOC or not. Both NTD NAbs lost all neutralization potency against B.1.351, while COVA2-17 retained neutralizing activity against P.1 (Fig. 3B). The 5-fold decrease in neutralization potency for COVA2-17 against B.1.1.7 is consistent with earlier findings (*32*). Strikingly, SARS-CoV cross-NAb COVA1-16 did not lose potency against any VOC, in line with previous results (*10*). The preservation of COVA1-16 potency (0.02 - 0.16 µg/mL for all VOC) as well as its remarkable breadth are consistent with the epitope of COVA1-16 remaining virtually unchanged throughout sarbecovirus evolution and after >1 year of global SARS-CoV-2 evolution (*29*), thereby underlining the importance of having such NAbs readily available as therapeutics against emerging SARS-CoV-2 VOC.

## Discussion

In this study, we show that COVID-19 convalescent patients as well as mRNA vaccine recipients sampled at the expected peak of their immunity, showed a marked decrease in binding and neutralization potency against two of three VOC currently circulating (Fig. 2, A and B). While patients who experienced mild infection have lower binding and neutralization titers against the original virus and often no activity against B.1.351 and P.1, all hospitalized convalescent patients and all but one vaccine recipient maintain neutralizing titers against the three VOC (Fig. 2C). Although the observed cross-reactivity is considerable, the substantial reduction in binding and neutralization titers against VOC B.1.351 and P.1 could possibly leave them more vulnerable to a (re-)infection with these VOC. Indeed, first reports of breakthrough infection in vaccine recipients are starting to emerge (*33*).

To date, most studies have focused on a specific VOC or on a specific set of samples (i.e. vaccine recipients, convalescent patients or NAbs) (*4, 10, 16, 20*). Here, through systematic comparison of non-hospitalized patients, hospitalized patients and vaccine recipients in the context of multiple VOC, we are able to assess the impact of specific sets of mutations. We observed that serum binding titers were impacted most by the P.1 VOC, while for neutralizing titers against B.1.351 showed the largest reduction in all three groups studied (Fig. 1C and 2B). This irregularity might be explained by non-neutralizing Abs targeting the S2 domain or NTD being more affected by the amino acid substitutions in P.1 compared to B.1.351. The most striking differences between B.1.1.7 and the other two VOC are the high number of mutations in the S2 domain and the lack of RBD mutations in B.1.1.7 that would result in a reduction in potency of RBD-targeting NAbs. Moreover, B.1.351 and P.1 show different neutralization signatures, most notably in hospitalized patients and vaccine recipients that have higher neutralization titers in general (Fig. 2B). While we and others have shown the E484K mutation impacts binding and neutralization by serum and RBD NAbs greatly (fig. S3B) (*34*), the discrepancy in neutralizing titers between B.1.351 and P.1 in vaccinees specifically might be because of the difference in amino acid substitution at position 417, as indicated by the NAb binding to the RBD variants. Alternatively, this discrepancy in neutralizing titers, despite similarity in their RBD, could be indicative of the NTD substitutions in B.1.351 causing a greater decrease in neutralizing titers than those found in P.1, which is reflected by NTD-targeting NAb COVA2-17. These results provide additional guidance for the risk evaluation of newly emerging variants during routine surveillance.

An implication of this study could be that (re-)infections with new VOC will become more prevalent, especially later on when antibody titers have declined. However, the question remains whether infection with an emerging, antigenically-altered variant will result in a less severe course of infection. While circulating neutralizing antibodies are expected to play a dominant role in preventing infection, memory B cells might prevent severe disease, as they quickly gear up to generate new neutralizing antibodies. Our finding that NAbs that have lost their neutralization capacity against VOC can often still bind to VOC S proteins, suggests that memory B cells of this specificity would still be able to recognize incoming VOC, and affinity-mature to regain neutralization capacity, resulting in faster virus clearance than during *de novo* immune responses. Indeed, the first results of the Ad26.CoV2.S phase 3 clinical trial indicate that the vaccine, coding for the WT S protein, does result in a milder disease course after infection with the B.1.351 VOC(*35*). It remains to be seen if this effect persists when antibody levels decline over time.

Some real-world consequences from the observations we made here can be gleaned from phase 3 efficacy studies with vaccines based on the Wuhan Hu-1 strain and performed in geographic areas where VOC dominated during the trial. First, the AstraZeneca ChAdOx1-S vaccine, which was tested in the United Kingdom when WT and B.1.1.7 were co-circulating, was subtly less effective against B.1.1.7 compared to the original virus (∼70% versus ∼81%) (*36, 37*), while it was virtually ineffective in South Africa where B.1.351 was dominant during the trial (*38*). The Janssen COVID-19 vaccine was tested in many countries including the United States and South Africa. The vaccine was 74% effective at preventing moderate-to-severe disease in the United States, where 97% of sequenced viruses were of the reference Wuhan-Hu-1 D614G strain, whereas in South Africa 95% of sequenced viruses were B.1.351 and vaccine efficacy fell to 52% (*35*). Similar trends were reported for the Novavax subunit vaccine in a press release.

Our results and those of others (*35, 36, 38*) strongly suggest that booster vaccines with antigenically diverse SARS-CoV-2 variants are needed to induce longer-lasting and more cross-reactive immunity. Indeed, multiple vaccine manufacturers have announced that they are already working on implementing VOC into their immunization regimen, with Moderna having started the evaluation of a booster vaccine candidate based on B.1.351 in a phase 1 clinical trial (*39*). Considering the timelines for modification of mRNA versus other vaccine platforms, the mRNA vaccines might offer advantages against the evolving virus.

We observed that NAb COVA1-16 retained activity against B.1.351 and P.1, both of which have the E484K mutation that has now also been observed in the B.1.1.7 lineage and multiple other lineages (*9, 12*). The intrinsic capacity of COVA1-16 to cross-neutralize SARS-CoV and its highly conserved epitope that remained unchanged throughout sarbecovirus evolution holds promise when considering SARS-CoV-2 evolution now and in the future. Moreover, COVA1-16 synergizes with another cross-reactive NAb CV38-142 to enhance neutralization of both SARS-CoV-2 and SARS-CoV (*40*). Together with the reduced effectiveness of several emergency-use authorized NAbs and NAb cocktails against B.1.351, our results re-emphasize the importance of not only having potent, but also cross-reactive NAbs available as a therapeutic tool(*10*). Therefore, particularly when the potency of COVA1-16 can be improved, it could be an attractive component of a pan-sarbecovirus NAb cocktail to prevent and treat infection with emerging SARS-CoV-2 variants and to have on the shelf for future sarbecovirus pandemics.

While our study aims to provide an overview of many different aspects of pre-existing immunity against VOC, there are some limitations. In this study, we present a vaccinee population that consists of primary health care workers that are generally of middle age, with only four (8%) participants being older than 60 (Table 1). However, we found no correlation between age and titers in convalescent patients against any of the VOC tested. Moreover, we have focused on the neutralization potential of immune sera and did not examine antibody effector functions which have been implicated previously in the control of SARS-CoV-2 infection (*41*). Similar to other studies, our convalescent and vaccinee sera samples were collected four to six weeks after infection and four weeks after vaccination (i.e. at the peak of immunity), respectively. While we present findings that may have implications for additional booster vaccines and the monitoring of VOC, we did not examine the aspect of waning antibody titers and how those might influence VOC binding and neutralization. Finally, we have merely examined an early line of defense against infection with SARS-CoV-2 (i.e. serum antibody levels), while we expect memory B cells to play an important part in re-infections with SARS-CoV-2 VOC. Indeed, swift re-activation of memory compartments may lead to reduced transmissibility, a milder course of disease and a more potent immune response.

In conclusion, we observed a substantial reduction in binding and neutralization potency against all three VOC in the majority of samples, with the individuals with lowest binding and neutralization titers losing potency altogether, especially to B.1.351 and P.1. These data have implications for the degree to which pre-existing immunity can protect against subsequent infection with VOC, the possible need of vaccine modifications in order to increase immune coverage and the use of monoclonal antibodies of therapeutics against SARS-CoV-2. An important outstanding question is what else the virus has in store for us and whether VOC, when not brought under control, will evolve further and continue to escape from humoral immunity induced by infection or vaccination.

## Materials & methods Study design

The sera of 69 SARS-CoV-2 infected adults were collected four to six weeks after symptom onset through the cross-sectional COVID-19 Specific Antibodies (COSCA) cohort (NL 73281.018.20) as described previously (*25*). All participants had at least one nasopharyngeal or oropharyngeal swab positive for SARS-CoV-2 as determined by quantitative reverse transcriptase-polymerase chain reaction (qRT-PCR, Roche LightCycler480, targeting the Envelope-gene 113bp). Participants were included from the start of the COVID-19 pandemic in the Netherlands in March 2020 until the end of January 2021. To investigate the humoral response to the Pfizer-BioNTech COVID-19 mRNA vaccine we used sera collected four weeks after the second dose of 50 health care workers who were included at the beginning of the pandemic in March 2020 in a prospective serologic surveillance cohort in two tertiary medical centers in the Netherlands (S3 cohort; NL 73478.029.20). Participants of the surveillance cohort were tested routinely for seroconversion by using a WANTAI (WS-1096) SARS-CoV-2 RBD total immunoglobulin serum enzyme-linked immunosorbent assay (ELISA) and underwent additional PCR testing when experiencing COVID-19 related symptoms. Participants were excluded when any of the above tests were indicative of a SARS-CoV-2 infection. Both studies were conducted at the Amsterdam University Medical Centers in the Netherlands and approved by the local ethical committee. All individuals included in this study gave written informed consent before participating.

### Protein design

The S constructs contained the following mutations compared to the WT variant (Wuhan Hu-1; GenBank: MN908947.3): deletion (Δ) of H69, V70 and Y144, N501Y, A570D, D614G, P681H, T716I, S982A and D1118H in B.1.1.7; L18F, D80A, D215G, L242H, R246I, K417N, E484K, N501Y, D614G and A701V in B.1.351; L18F, T20N, P26S, D138Y, R190S, K417T, E484K, N501Y, D614G, H655Y and T1027I in P.1. They were ordered as gBlock gene fragments (Integrated DNA Technologies) and cloned PstI/NotI in a pPPI4 expression vector containing a hexahistidine (his) tag with Gibson Assembly (ThermoFisher). All S constructs were verified by Sanger sequencing and subsequently produced in HEK293F cells (ThermoFisher) and purified as previously described (*25*). The RBD constructs contained the following mutations: N501Y in B.1.1.7; K417N, E484K and N501Y in B.1.351; K417T, E484K and N501Y in P.1; E484K as a single mutant. They were made by introducing the mutations into the SARS-CoV-2 RBD (331-528 amino acids)-StrepII construct, kindly provided by Dr. Premkumar Lakshamanane (*42*), using a QuikChange site-directed mutagenesis kit (Agilent Technologies). All RBD constructs were produced in ExpiCHO cells and purified as previously described (*43*).

### Multiplex protein microarray

HCoV-PMA slides were produced as described previously (*23*). WT and VOC (B.1.1.7, B.1.351 and P.1) S proteins were spotted in duplicate in three drops of 333 pL each on 24-pads nitrocellulose-coated slides (ONCYTE AVID, GraceBio Labs, Bend, USA) by using a non-contact Marathon Arrayjet microarray spotter (Roslin, UK). Printed micro-array slides were pre-treated with Blotto blocking buffer (ThermoFisher) to avoid non-specific binding. Sera were tested in four 4-fold dilutions starting at 1:20, diluted in Blotto buffer containing 0.1% Surfact-Amps20 (ThermoFisher) as previously described (*23*). Subsequently, slides were incubated with goat anti-human IgG, F(ab’)2 fragment specific, Alexa Fluor 647-conjugated (Jackson Immunoresearch, West Grove, USA), diluted 1:1000 in Blotto buffer with 0.1% Surfact-Amps20 as described. Incubation steps were followed by a washing step with 1× phosphate-buffered saline (PBS) with 0.1% Tween. After the last wash, slides were washed with sterile water and dried. Day-to-day variations were monitored by including a SARS-CoV-2 positive control serum in each test round. An in-house SARS-CoV-2 standard was included in each test batch to correct for test-to-test variation. If the titer of the positive control deviated more than 2-fold from the expected titer, the test batch was rejected and repeated.

### Pseudovirus design

The B.1.1.7, B.1.351 and P.1 pseudovirus constructs contained the same mutations as the S constructs. They were ordered as gBlock gene fragments (Integrated DNA Technologies) and cloned SacI/ApaI in the pCR3 SARS-CoV-2-S_Δ19_ expression plasmid (GenBank: MT449663.1) (*44*) using Gibson Assembly (ThermoFisher). The D614G pseudovirus construct was made using QuickChange Site-Directed Mutagenesis (Agilent Technologies). All constructs were verified by Sanger sequencing. Pseudovirus was produced by co-transfecting the pCR3 SARS-CoV-2-S_Δ19_ expression plasmid with the pHIV-1_NL43_ ΔEnv-NanoLuc reporter virus plasmid in HEK293T cells (ATCC, CRL-11268) (*44, 45*). Cell supernatant containing the pseudovirus was harvested 48 h post transfection and stored at −80°C until further use.

### Pseudovirus neutralization assay

HEK293T/ACE2 cells kindly provided by Dr. Paul Bieniasz (*44*) were seeded at a density of 20,000 cells/well in a 96-well plate coated with 50 μg/mL poly-L-lysine 1 day prior to the start of the neutralization assay. NAbs (1-50 μg/mL) or heat-inactivated sera samples (1:100 dilution) were serial diluted in 5-fold resp. 3-fold steps in cell culture medium (DMEM (Gibco), supplemented with 10% FBS, penicillin (100 U/mL), streptomycin (100 μg/mL) and GlutaMax (Gibco)), mixed in a 1:1 ratio with pseudovirus and incubated for 1 h at 37°C. These mixtures were then added to the cells in a 1:1 ratio and incubated for 48 h at 37°C, followed by a PBS wash and lysis buffer added. The luciferase activity in cell lysates was measured using the Nano-Glo Luciferase Assay System (Promega) and GloMax system (Turner BioSystems). Relative luminescence units (RLU) were normalized to the positive control wells where cells were infected with pseudovirus in the absence of NAbs or sera. The inhibitory concentration (IC_50_) and neutralization titers (ID_50_) were determined as the NAb concentration and serum dilution at which infectivity was inhibited by 50%, respectively using a non-linear regression curve fit (GraphPad Prism software version 8.3) (*45*).

### Authentic virus neutralization assay

SARS-CoV-2 virus neutralization assays were performed as described previously (*46, 47*). Duplicates of 2-fold serial dilutions (starting at 1:10) of heat-inactivated sera (30 m, 56°C) were incubated with 100 median tissue culture infectious dose (TCID_50_) of SARS-CoV-2 strains hCoV-19/Netherlands/ZuidHolland_10004/2020, D614G (WT) and hCoV-19/Netherlands/ NoordHolland_10159/2021 (B.1.351, EVAg cat.nr. 014V-04058) at 35°C for 1 h in 96-well plates. Vero-E6 cells were added in a concentration of 20,000 cells/well and incubated for 72 h at 35°C. The serum virus neutralization titer (VNT_50_) was defined as the reciprocal value of the sample dilution that showed a 50% protection of virus growth. Samples with titers ≥ 20 were defined as SARS-CoV-2 seropositive.

### Monoclonal antibodies

The NAbs used in this study were isolated from participants in the “COVID-19 Specific Antibodies” (COSCA) study as described before. NAbs were produced in HEK293F (ThermoFisher) cells as previously described (*25*).

### Bio-layer interferometry (BLI)

BLI experiments were performed on an Octet K2 (ForteBio). Ni-NTA biosensors (ForteBio) were loaded with 20 µg/mL of each of the four different his-tagged S proteins (WT, B.1.1.7, B.1.351 and P.1) in running buffer (PBS, 0.02% Tween-20, 0.1% BSA) for 300 s. After the biosensors were washed in a well containing running buffer to remove excess protein, the biosensors were dipped in a well containing 30 µg/mL NAb in running buffer for 120 s to measure association. Next, the biosensors were moved to a well containing running buffer for 120 s to measure dissociation of the S protein-NAb complexes.

### Enzyme-linked immunosorbent assay (ELISA)

Purified RBD-StrepII proteins (200 ng/well) were coated overnight onto Nunc MaxiSorp 96-well plates (Thermo Fisher) at 4ºC. The plates were washed three times with PBS/0.05% Tween-20 before blocking with 5% milk/PBS for 1 h at RT. After washing as described above, COVA NAbs, serially diluted in 2% milk/PBS, were added and incubated for 1 h at RT. Bound NAbs were detected using horseradish peroxidase (HRP)-conjugated goat anti-human IgG (1:3000) in 2% milk/PBS for 1 h at RT. After washing, the color reaction was developed using 3,3′,5,5′-tetramethylbenzidine (TMB) substrate (Thermo Scientific). The reaction was stopped by adding 0.3 N sulfuric acid and OD_450_ was read using an Enspire instrument (Perkin Elmer).

### Visualization and statistical analysis

Data visualization and statistical analyses were performed in GraphPad Prism software (version 8.3). The non-parametric Friedman test was performed to assess statistical differences for paired samples, while the non-parametric Kruskal-Wallis test was performed for unpaired samples. Significance is denoted as ****, *p* < 0.0001; ***, *p* < 0.001; **, *p* < 0.01; *, *p* < 0.05; ns, not significant.

## Data Availability

All data are available in the main text or the supplementary materials. Reagents used in this study are available upon reasonable request under an MTA with Amsterdam UMC.

## Acknowledgments

We thank Dr. Paul Bieniasz for donating cells and reagents for pseudovirus neutralization assays; Johan Reimerink, Fion Brouwer, Marieke Hoogerwerf and Tarek Munawar for technical assistance; Bas J. Verkaik, Orlane J.A. Figaroa, Peter J. de Vries, Tessel M. Boertien and Maria Prins for support of the COSCA study and all the participants of the COSCA and S3/HCW studies.

## Funding

Netherlands Organization for Scientific Research (NWO) Vici grant (RWS)

Bill & Melinda Gates Foundation, Collaboration for AIDS Vaccine Discovery (CAVD) grant INV-002022 (RWS)

Amsterdam UMC AMC Fellowship (MJvG)

Bill & Melinda Gates Foundation, COVID-19 Wave 2 mAbs grant INV-024617 (MJvG)

Fondation Dormeur, Vaduz (MJvG, RWS)

HIVRAD grant P01 AI 110657 (JPM)

R01 AI 36082 (JPM)

Netherlands Organization for Health Research and Development ZonMw & the Amsterdam UMC Corona Research Fund (Amsterdam UMC COVID-19 S3/HCW study group)

## Author contributions

Conceptualization: TGC, IB, MJvG, DE, RWS

Funding acquisition: MJvG, RWS

Investigation: TGC, IB, MP, JAB, MO, GG, CV, RM, DG, JLS, DNC, ICM, TM,

Methodology: TGC, IB, MP, JB, MJvG, DE, RWS

Project administration: TGC, IB, MJvG, DE, RWS

Resources: KvdS, BA, AHAL, HDGW, EW, Amsterdam UMC COVID-19 S3/HCW study group, JJS, MKB, GJdB

Supervision: JPM, MDJ, CR, MJvG, DE, RWS

Writing – original draft: TGC, IB, KvdS, MJvG, DE, RWS

Writing – review & editing: all authors

